# Time-Varying Coactivation Pattern and Knee Joint Kinematics and Kinetics Following ACL Reconstruction: A Statistical Parametric Mapping Approach

**DOI:** 10.64898/2026.09.30.26364410

**Authors:** Imaad Virani, Mohsen Alighanbari, Timothy Lowe

## Abstract

**Introduction:** Knee muscle coactivation is elevated after anterior cruciate ligament reconstruction (ACLR), contributing to altered joint loading and post-traumatic osteoarthritis. Existing methods average coactivation within predefined gait phases and require priori designation of muscles roles, limiting the resolution for when differences occur.

**Objective:** Characterize knee joint coactivation and dynamics throughout gait in individuals with ACLR using statistical parametric mapping.

**Methods:** Ten individuals with unilateral ACLR and ten healthy matched controls performed overground walking at their preferred speed, while kinematics, kinetics, and electromyography of the quadriceps, hamstring, and gastrocnemius muscles were recorded. Coactivation was quantified using time-varying, multi-muscle coactivation, and was compared using a one-tailed statistical parametric mapping (SPM) t-test. Knee joint angle, angular speed, and moment were compared using SPM Hotelling’s T^2^ tests with Bonferroni-adjusted post hoc comparisons (*α* = 0.05).

**Results:** Coactivation was greater in the ACLR group at 0.01%-6.14%, 35.23%-40.94%, 69.73-71.99%, 72.12-77.31%, 77.89-78.05%, and 94.63%-95.87% (p < 0.0001) of stride. The ACLR group also demonstrated greater knee internal–external rotation angle at 0.01%-47.34%, 52.14%-61.87%, and 66.47%-100% (p <0.0001) and lower knee flexion angle at 55.79%-65.93% (p = 0.004) of stride, than healthy controls. Finally, a smaller knee flexion–extension moment was observed in the ACLR group at 57.43%-62.18% (p = 0.005) of stride.

**Conclusions:** Coactivation after ACLR is elevated at loading response, late mid-stance, terminal stance and early push-off phase. Greater coactivation was observed during weight-acceptance phases, whereas lower knee moments characterized initial swing. These alterations are localized rather than uniformly distributed, supporting phase-targeted rehabilitation.

## 1. Introduction

Approximately 400,000 ACL related injuries occur each year, with the highest incidence occurring in sports participants. ^1,2^ ACL tears result in severe functional impairments, especially in pivoting motions, increased joint laxity, and are associated with more frequent and earlier incidence of other knee surgeries. ^3,4^ This is a rapidly growing public health issue as the incidence of ACL injuries has had the largest growth rate per year among knee injuries. ^5^ Furthermore, they contribute to an additional 30,000-38,000 patients with knee osteoarthritis and an additional 25,000-30,000 total knee arthroplasties each year. ^6^

The mainstay of treatment for this injury is ACL reconstruction (ACLR) surgery. Patient-reported outcomes, muscle strength, and gait mechanics have all been shown to improve in the first 12 months post-reconstruction. ^7^ However, muscle coactivation between the quadriceps and hamstrings is increased in relation to both the contralateral leg and healthy controls ^8–10^. Muscle coactivation refers to the simultaneous activation of agonist and antagonist muscle groups, which has been proposed to reflect a deliberate neural control strategy. ^11–13^ During gait, knee joint coactivation refers to concurrent activity of knee flexors and extensors, intended to increase active joint stability, often at the expense of strength, flexibility, and cartilage health. ^9,14,15^ An increase in knee coactivation can be triggered by multiple phenomena, including but not limited to joint inflammation, arthrogenic muscle inhibition (AMI), and protective neuromuscular alterations. ^16,17^ AMI in particular, involves pre-synaptic modulation of input to motor neurons, which reduces voluntary activation of that muscle. ^18^ Both the ACL injury itself and the subsequent ACLR cause trauma to the joint, producing inflammation that is considered to trigger reflexive inhibition of the quadriceps, which is associated with reduced activation and muscle atrophy. ^16,19^

The current literature suggests that the increased activation of hamstrings post-ACLR is a deliberate mechanism to assist with knee joint stability, and it further contributes to the increased levels of coactivation seen in these individuals. ^9,20^ In the early post-op period, this response is generally considered protective. ^20,21^ It enhances dynamic knee stability through increased hamstring contribution, which has been associated with reduced risk of secondary injury. ^21–23^

However, when coactivation persists beyond this acute window, it may begin to exert a degradative effect on the knee joint. These biomechanical changes have been observed in a study of quadriceps function following ACLR, in which participants with ACLR demonstrated lower quadriceps peak torque (1.86 versus 2.56 Nm/kg), rate of torque development (39.4 versus 52.9 Nm/s/kg), and EMG amplitude (0.25 versus 0.37 mV), along with greater hamstrings co-activation (27.2% versus 14.3%) compared to healthy controls. ^9^ Similar findings have been reported during isometric knee extension, where hamstrings/quadriceps coactivation was 15.1% compared to 7.5% in controls. ^10^

While the progression of muscle coactivation has not been directly characterized in long-term studies, persistent deficits in knee extensor and flexor strength at one-year post-ACLR indicate that neuromuscular impairments do not fully resolve in that time. ^24,25^ Coactivation has also been associated with stiffer gait patterns, including reduced knee flexion displacement, altered sagittal-plane knee moments, increased vertical ground reaction force loading rate, and proximal redistribution of mechanical work across the lower extremity. ^8,26,27^ Because reduced knee flexion limits the capacity of the quadriceps to attenuate impact loads through eccentric work, this movement pattern may increase mechanical loading of passive structures such as the cartilage and meniscus, consistent with findings from landing tasks. ^15^ Over time, this altered loading distribution has also been linked to cartilage degeneration, predisposing these individuals to knee osteoarthritis. ^15,28,29^

Recent research has attempted to characterize temporal patterns of coactivation throughout the gait cycle to better understand the mechanical pathogenesis of osteoarthritis and inform potential modifications to clinical management. ^8^ However, despite the increasing association between coactivation and adverse outcomes following ACLR, establishing a causal relationship has proven difficult. This challenge is partly attributable to substantial methodological variability in how coactivation is quantified. Traditional approaches rely on a priori designation of muscles as agonists and antagonists and calculate a co-contraction index to quantify the relative level of muscle activity surrounding a joint. ^13^ The most common approach uses normalized surface electromyography (EMG) data; however, considerable variation exists in how these data are processed and combined to derive coactivation measures. ^12,13,30^ Other studies have used moment-based indices or more recent approaches, such as wavelet analysis, to characterize muscle coactivation. ^31,32^

Although each of these approaches has its merits, the inability to account for all muscles contributing to joint coactivation, the requirement to a priori designate muscle role, and the reliance on phase-based analysis limit the comprehensive and temporally localized characterization of joint coactivation during gait. Some studies utilized medial and lateral designation of musculature while other studies solely focused on a two-muscle model of the quadriceps and hamstrings, rendering them fundamentally incomparable. ^8,9^ The a priori designation of large muscle groups as agonists and antagonists inherently limits the assessment of knee joint coactivation. ^13^ The potential effects from the many other muscles associated with movement of the knee joint are obscured, and, because muscle roles shift continuously during gait, ratio-based indices that depend on this assignment can consequently return values exceeding 100%, conflicting with the definition of coactivation. Finally, studies that have explored improving the time resolution of coactivation throughout the entire gait cycle have analyzed the cycle in discrete phases rather than performing a continuous analysis. ^8^ As a result, the combination of the limitations of previous methods and the heterogeneity between them makes it difficult to compare time-resolved coactivation findings in this population.

Coactivation also shapes the mechanical behavior of the knee during gait. In ACLR limbs, greater coactivation has been associated with reduced knee flexion displacement, smaller peak internal knee extension moments, and greater peak internal knee valgus moments during walking. ^8^ Comparable relationships have been observed during landing, where individuals with ACLR demonstrated greater coactivation, less knee flexion, and elevated tibiofemoral compressive forces relative to controls. ^15^ Interventions that acutely reduce hamstring coactivation produce a reversal of these mechanical changes, increasing knee flexion and knee extension moment while lowering compressive force, limb stiffness, and loading rate. ^27,28^ Altered knee angle and moment profiles have themselves been reported at multiple points during stance following ACLR and have been linked to the development of medial compartment osteoarthritis. ^29,33,34^ Because the mechanical consequences of altered neuromuscular control are expressed in joint angle and moment, characterizing coactivation alone provides an incomplete account of the reconstructed knee.

Therefore, the present study has two aims. The first was to assess global knee joint coactivation continuously across the gait cycle in individuals with ACLR and healthy controls, using a multi-muscle model that requires no a priori designation of muscle role, to determine precisely when coactivation patterns differ between groups. The second was to perform a time-localized assessment of knee joint angle and moment in the same participants, to identify when the mechanical behavior of the reconstructed knee diverges from that of healthy controls. We expected that this approach would resolve differences in both coactivation and joint mechanics that phase-averaged analyses cannot detect, capturing quantitative differences in magnitude as well as qualitative differences in muscle coordination and recruitment during gait.

## 2. Method

For the present study, 10 participants with unilateral ACLR (Age: 26.6 ± 1.9 yrs., Height: 167.4 ± 0.1 cm, Weight: 73 ± 12.7 Kg) and 10 healthy young adults (Age: 26.9 ± 1.3 yrs., Height: 167.6 ± 0.09 cm, Weight: 61.6 ± 9.7 Kg) were recruited. The study protocol is in accordance with the Declaration of Helsinki and has been approved by the University of Texas at Austin Institutional Review Board (IRB). All participants were familiarized with the data collection and signed a written consent prior to data collection. Kinematic data was recorded using 12 Vicon cameras (Vicon Motion Systems Ltd., UK) at 100Hz with full-body plug-in gait marker placement. Kinetic data was recorded at a sampling frequency of 2000Hz (Bertec Corp.). Muscle activity was recorded using Delsys wireless surface electromyography (sEMG) at 2000Hz (Delsys Inc.). The sEMG was recorded unilaterally from the vastus lateralis (VL), vastus medius (VM), rectus femoris (RF), semitendinosus (ST), biceps femoris (BF), and medial (GM) and lateral gastrocnemius from the ACLR limb (3 Left and 7 right legs) and the matched control limb. Electrode attachment and skin preparation were performed according to the SENIAM guideline.

Participants performed a 5-min warmup exercise on a cycle ergometer, ^10^ then performed overground walking at a comfortable, self-selected speed (fig. 1). Heel strike was defined as vertical ground force (vGRF) > 20 N, the best three strides were selected for further analysis. Kinematic data were filtered with the 4th-order zero-lag low-pass Butterworth filter with a cut-off frequency of 6Hz. Joint moments were then normalized to height and weight.

**Figure 1:**
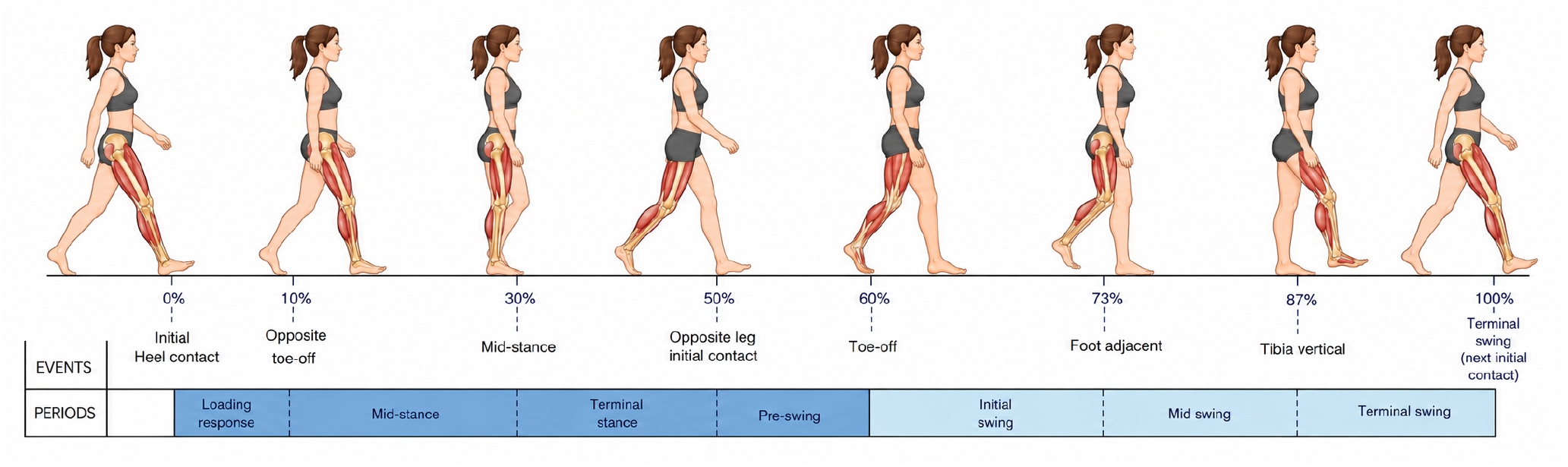
The cycle is normalized from initial heel contact of the reference limb (0%) to its subsequent initial contact (100%). Stance phase (approximately 0–60%) includes loading response, mid-stance, and terminal stance. Swing phase (approximately 60–100%) includes initial, mid-, and terminal swing. Dark blue indicates the stance phase and the light blue indicates swing phase.

The sEMG data were filtered using a 5th-order band-pass zero-lag Butterworth filter with a cut-off frequency of 10-500Hz. The sEMG data were then linear enveloped using a 2nd-order low-pass zero-lag Butterworth filter with a cutoff frequency of 25Hz and normalized to the peak muscle activity during walking trials. The normalized EMG was used to calculate the time-varying multi-muscle coactivation function (TMCf) for the knee joint.

### 2.1 TMCf

The TMCf, a time-varying multi-muscle coactivation method, can be calculated with eq. (1). ^13^

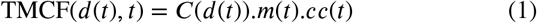

Where the *C*(*d*(*t*)) is a sigmoid weight reduction coefficient ranging between 0 and 1, the *m*(*t*) is the mean behavior of all the EMGs, and the *cc*(*t*) is the correction coefficient to control the TMCf behavior in the case that only two muscles were used. The sigmoid weight-reduction coefficient, which accounts for the mean differences between each pair within the exponent function, can be calculated using eq. (2). ^35^

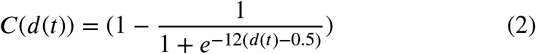

Where the e is Euler’s constant serving as a nonlinear mapping tool, and the *d*(*t*) is the mean differences between each pair of muscle EMG samples at time t, which can mathematically be expressed as eq. (3).

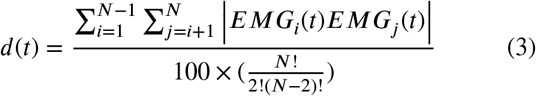

Where the 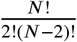 is the total number of possible differences between each muscle pair’s *EMG*_*i*_(*t*). The mean behavior of all the EMGs in eq. (1) (*m*(*t*)) is derived from the Rudolph function, ^36^ which can be calculated with eq. (4). ^13^

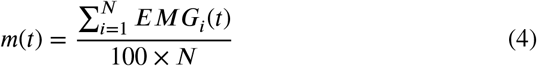

Where the *EMG*_*i*_(*t*) is the amplitude of the normalized EMG at time t, and N is the number of muscles in the TMCf calculation. Finally, a correction coefficient to control the TMCf behavior in case of two muscle (*cc*(*t*)) were calculated with eq. (5).

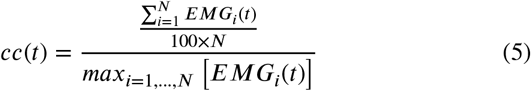

Where the *EMG*_*i*_(*t*) is the normalized EMG amplitu N is the number of EMG channels, and t is the sample time. The TMCf yields a scalar signal of multi-muscle coactivations for the knee joint. The knee joint TMCf calculated with VL, VM, RF, ST, BF, GM, and GL muscl All the data analysis was performed in MATLAB R20 (The MathWorks, Inc., Natick, MA, USA).

### 2.2 Knee joint dynamics

The knee joint angle and moment in the sagittal, frontal, and transverse planes were exported from Vicon. Joint angle and moment were then filtered with a fourth-order zero-lag low-pass Butterworth filter with a cut-off frequency of 6Hz. The joint moment was normalized to body weight and height and used for statistical analysis. Finally, the joint angular instantaneous speed was calculated using the three-point finite-difference formula and used for statistical analysis.

### 2.3 Center of Mass

The center of mass (CoM) trajectories were exported from Vicon. The CoM trajectories were then filtered with a low-pass zero-lag Butterworth filter with a cut-off frequency of 6 Hz. Each CoM trajectory was then normalized to the CoM initial distance and time-normalized to 10000 time points. The instantaneous CoM resultant distances were then calculated using the 3-dimensional Euclidean distance formula. The resultant CoM distance was then normalized to the participant’s height. Finally, the instantaneous CoM speed was calculated using the three-point formula as is used for statistical analysis.

### 2.4 Statistical analysis

An SPM t-test was used to assess differences in CoM distance and instantaneous speed between the ACLR and control groups. A one-tailed unpaired SPM t-test assessed the difference in TMCf of the knee joint between ACLR and control groups. SPM Hotelling *T*^2^ test was used to assess the differences in knee joint angle, angular speed, and moment between the ACLR and control groups. An SPM t-test with adjusted Bonferroni test was then used for the post hoc test. Group comparisons were performed using statistical parametric mapping (SPM) with SPM1d version 0.4.1 (https://spm1d.org/). ^37^ All statistical analyses were performed in MATLAB with a significance level set to *α* < 0.05.

## 3. Results

The SPM t-test analysis for resultant CoM distance and resultant CoM instantaneous speed showed no statistical difference between the ACLR and health control group (fig. 2). Knee coactivation patterns between the ACLR and Control group indicated that ACLR group had a greater coactivation at 0.01% to 6.14% (|*t*\*| = 4.01, *t*_*max*_ = 5.7, *p* < 0.0001, *d*_*range*_ = 1.87 − 2.67), 6.95% to 7.62% (*t*\* = 3.96, *t*_*max*_ = 4.44, *p* = 0.0341, *d*_*range*_ = 1.84−2.05), 28.56% to 29.73% (*t*\* = 3.94, *t*_*max*_ = 4.67, *p* = 0.0152, *d*_*range*_ = 1.83 − 2.2), and 35.23% to 40.94% (*t*\* = 3.98, *t*_*max*_ = 6.41, *p* < 0.0001, *d*_*range*_ = 1.85 − 3.02) of stride cycle (fig. 3). The ACLR group had greater knee joint coactivation than the control group at 69.73% to 71.99 (*t*\* = 3.98, *t*_*max*_ = 4.3, *p* = 0.0005, *d*_*range*_ = 1.87 − 2.02), 72.12% to 77.31% (*t*\* = 3.98, *t*_*max*_ = 5.26, *p* < 0.0001, *d*_*range*_ = 1.88 − 2.48), 77.89% to 78.05% (*t*\* = 3.99, *t*_*max*_ = 4, *p* = 0.0489, *d*_*range*_ = 1.88 − 1.89), 90.51% to 91.13% (*t*\* = 3.98, *t*_*max*_ = 4.1, *p* = 0.0361, *d*_*range*_ = 1.87 − 1.91), 94.63% to 95.87% (*t*\* = 3.97, *t*_*max*_ = 5, *p* = 0.0136, *d*_*range*_ = 1.87 − 2.36), 98.43% to 98.86% (*t*\* = 3.97, *t*_*max*_ = 4.25, *p* = 0.0427, *d*_*range*_ = 1.87 − 2) of stride cycle.

**Figure 2:**
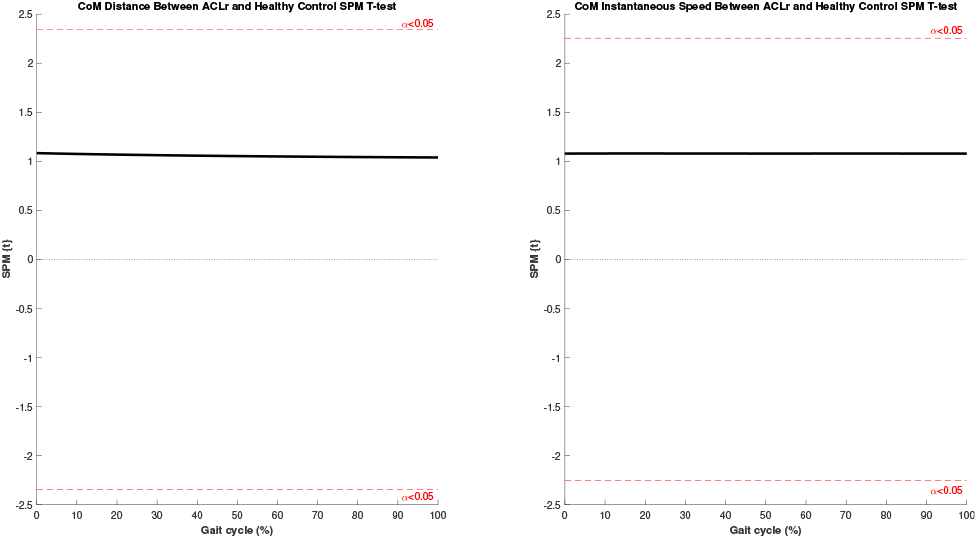
Similar Center-of-Mass Displacement and Instantaneous Speed Between ACLR and Control Groups. Statistical parametric mapping (SPM) analysis of resultant center-of-mass (CoM) displacement (left) and instantaneous speed (right) during walking. The solid black line represents the SPM t-statistic, and the dashed red line represents the statistical significance threshold. No significant between-group differences were identified, indicating comparable CoM displacement and walking speed between the ACLR and control groups.

**Figure 3:**
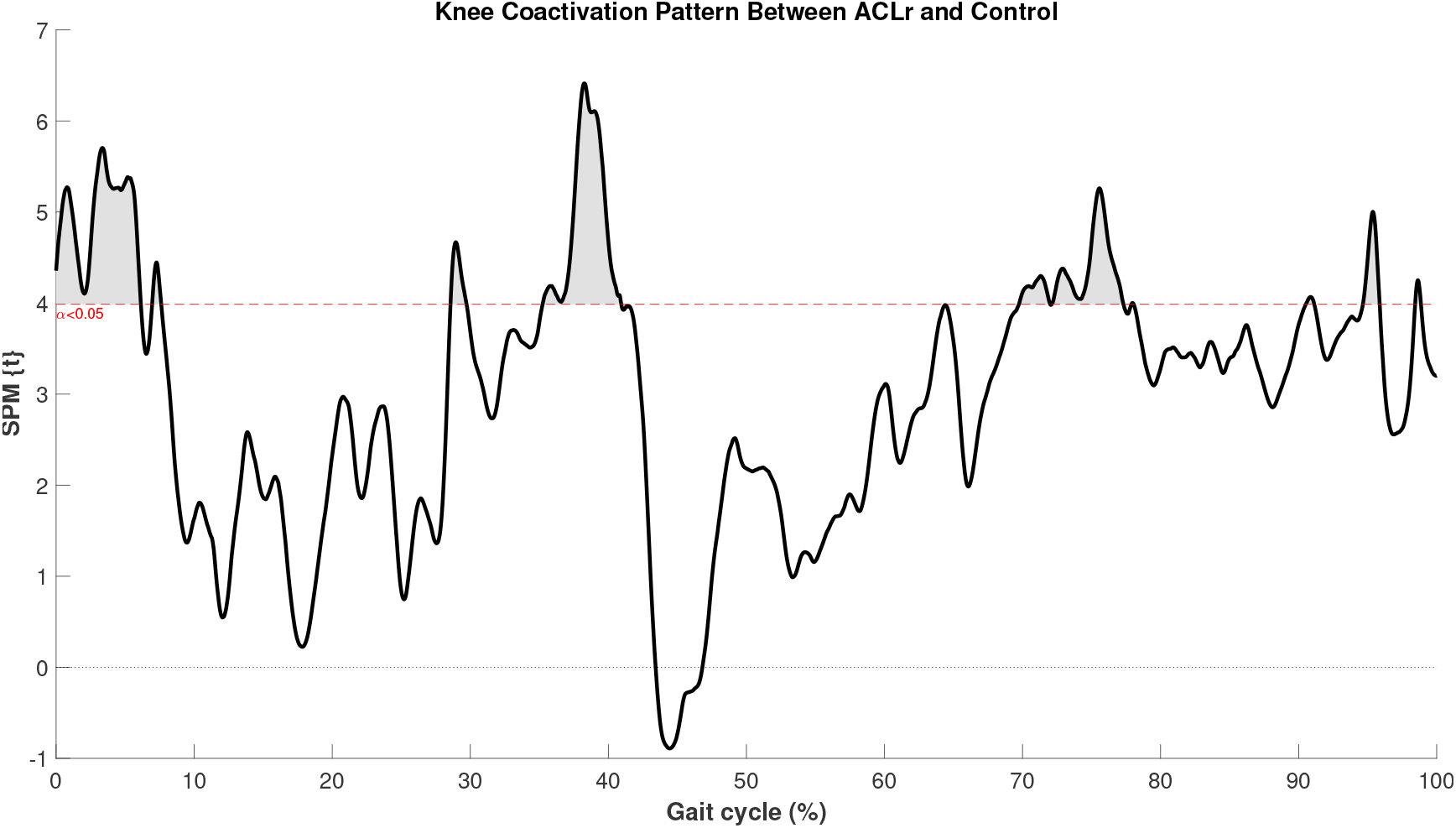
Greater Time-Varying Multi-Muscle Coactivation in the ACLR Limb During Specific Phases of the Gait Cycle. Statistical parametric mapping (SPM) analysis of time-varying, multi-muscle coactivation function (TMCf) across the gait cycle. The x-axis represents the gait cycle, time-normalized from heel strike to the subsequent ipsilateral heel strike and expressed as 0–100% of the stride. The y-axis represents the SPM{t} test statistic, a unitless value calculated at each point of the normalized gait cycle from the TMCf, which ranges from 0 to 100 and was derived from seven recorded knee muscles. The solid black line represents the SPM{t} trajectory, and the red dashed line represents the statistical significance threshold. Shaded clusters above the threshold indicate portions of the gait cycle in which coactivation differed significantly between groups, with cluster width representing the duration of the significant difference. Group differences were evaluated using a one-tailed independent-samples SPM t-test at *α* = 0.05. Coactivation was greater in the ACLR limb than in the control group within all significant clusters.

The SPM Hotelling *T*^2^ results indicated a significant difference in knee joint angle between ACLR and the control group during the overall stride cycle (*t*\* = 17.79, *t*_*max*_ = 53.05, *p* < 0.0001). Post hoc analysis indicated that the ACLR had a greater knee internal–external rotation angle at 0.01% to 47.34% (*t*\* = 3.22, *t*_*max*_ = 5.92, *p* < 0.0001, *d*_*range*_ = 0.84 − 1.53), 52.14% to 61.87% (*t*\* = 3.22, *t*_*max*_ = 3.42, *p* = 0.011, *d*_*range*_ = 0.84 − 0.89), and 66.47% to 100% (*t*\* = 3.24, *t*_*max*_ = 5.98, *p* = 0.0002, *d*_*range*_ = 0.84 − 1.55) of the stride cycle (fig. 4). The ACLR group also showed a smaller knee flexion angle at 55.79% to 65.93% (*t*\* = −3.4, *t*_*max*_ = −3.95, *p* = 0.004, *d*_*range*_ = −0.88*to*−1.02) of the stride cycle. However, no statistically significant difference was found for knee adduction–abduction angle.

**Figure 4:**
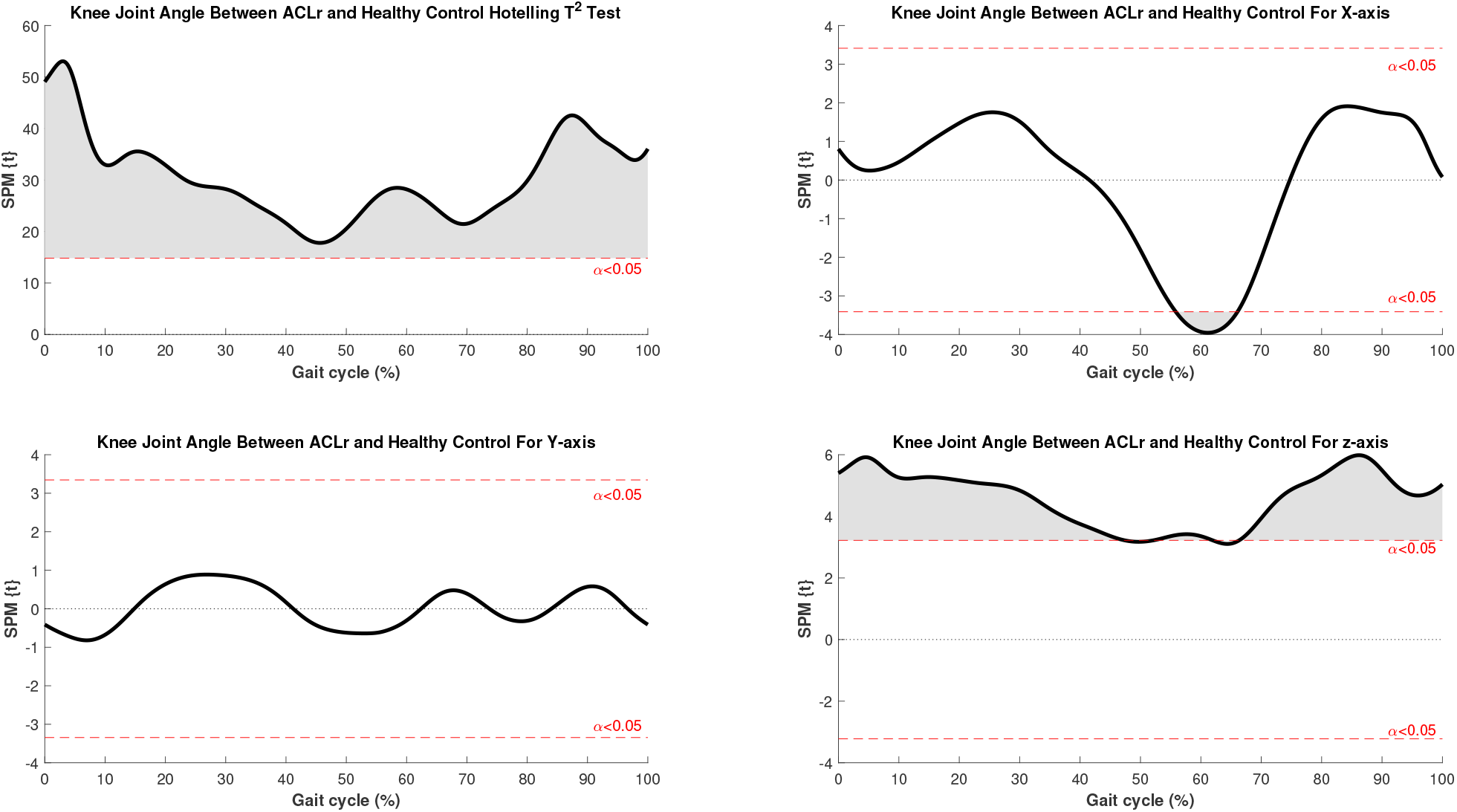
Altered Three-Dimensional Knee Joint Angle in the ACLR Limb Across the Gait Cycle. Statistical parametric mapping (SPM) analysis of knee joint angle. The upper-left panel presents the SPM{*T*^2^} trajectory from a Hotelling’s *T*^2^ test evaluating the three-dimensional knee joint angle simultaneously. The remaining panels present SPM{t} trajectories from post hoc analyses of the sagittal (flexion–extension, X), frontal (adduction–abduction, Y), and transverse (internal–external rotation, Z) components, with Bonferroni-adjusted significance thresholds. Joint angle was expressed in degrees. Clusters above the upper threshold indicate values greater in the ACLR limb, whereas clusters below the lower threshold indicate values lower in the ACLR limb. Knee joint angle differed between groups across much of the gait cycle, with greater internal–external rotation and lower flexion–extension angles in the ACLR limb; no significant between-group differences were identified in the adduction–abduction angle.

The SPM Hoteling *T*^2^ test results for knee joint angular speed indicated a significant difference between ACLR and the control group at 0.01% to 11.66% (|*t*\*| = 14.43, *t*_*max*_ = 19.82, *p* = 0.01), 25.61% to 72.42% (|*t*\*| = 14.42, *t*_*max*_ = 22.54, *p* = 0.026), 84.6% to 88.38% (|*t*\*| = 14.42, *t*_*max*_ = 14.71, *p* = 0.016), 93.93% to 100% (*t*\* = 14.44, *t*_*max*_ = 18.52, *p* = 0.024) of stride cycle. Post hoc analysis indicated that the ACLR group had greater knee adduction–abduction angular speed at 0% to 77.9% (*t*\* = 3.23, *t*_*max*_ = 4.5, *p* < 0.0001, *d*_*range*_ = 0.84 − 1.17) and 82.52% to 100% (*t*\* = 3.23, *t*_*max*_ = 4.29, *p* = 0.005, *d*_*range*_ = 0.84 − 1.11) of the stride cycle (fig. 5). Also, the ACLR group had greater internal–external knee rotation angular speed than the control group at 0.01% to 15.79% (*t*\* = 3.23, *t*_*max*_ = 4.28, *p* = 0.006, *d*_*range*_ = 0.84 − 1.11) and 89.32% to 100% of the stride cycle (*t*\* = 3.23, *t*_*max*_ = 3.98, *p* = 0.01, *d*_*range*_ = 0.84 − 1.03). However, no statistically significant difference was found between the ACLR and the control group for knee flexion–extension angular speed.

**Figure 5:**
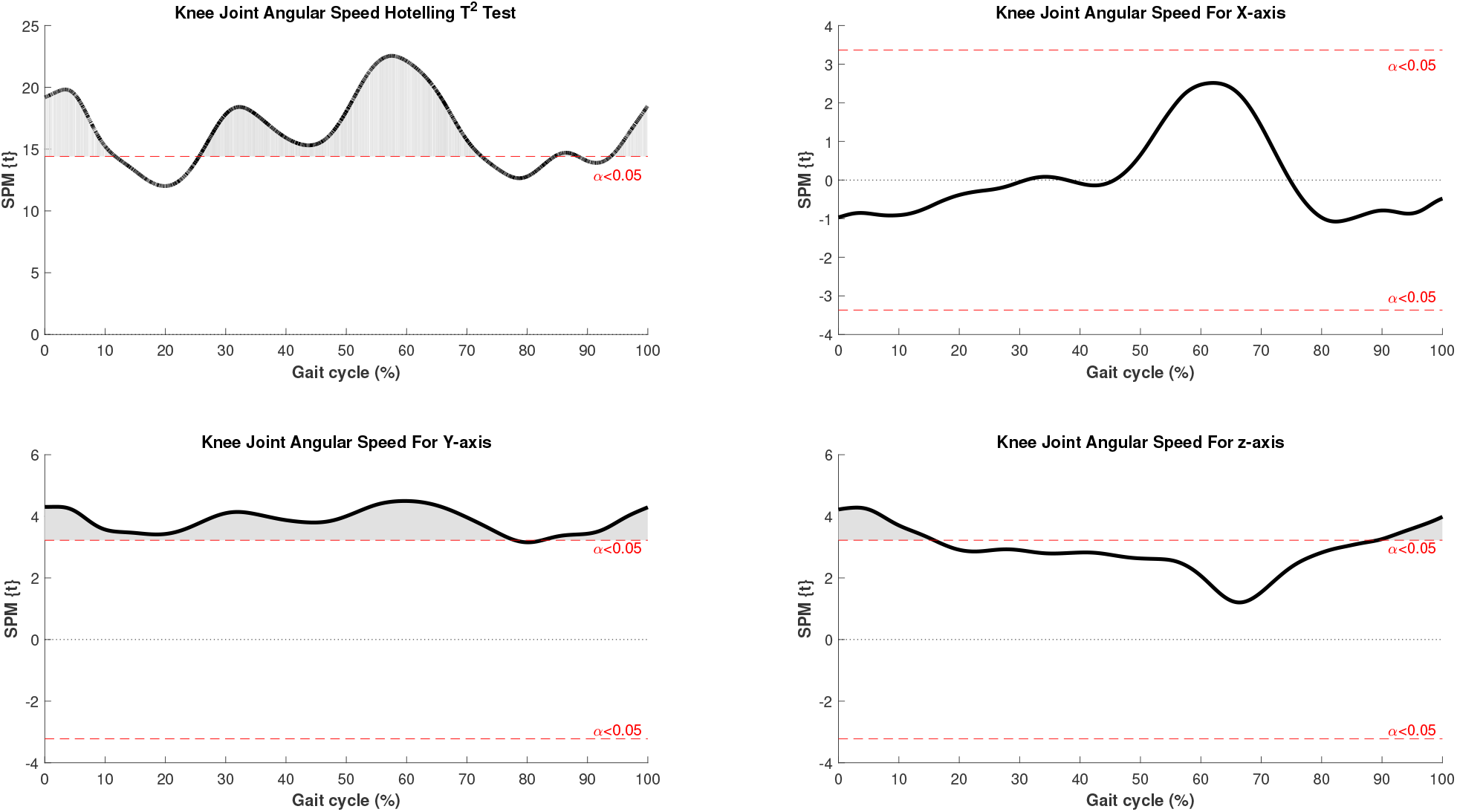
Greater Knee Joint Angular Speed in the ACLR Limb for Frontal and Transverse Plan Motion. Statistical parametric mapping (SPM) analysis of knee joint angular speed. The upper-left panel presents the SPM{*T*^2^} trajectory from a Hotelling’s *T*^2^ test evaluating the three-dimensional knee joint angular speed simultaneously. The remaining panels present post hoc SPM{t} analyses of the sagittal (flexion–extension, X), frontal (adduction–abduction, Y), and transverse (internal–external rotation, Z) components. Angular speed was calculated from the knee joint angle using a three-point finite-difference method and expressed in degrees per second. Angular speed was greater in the ACLR limb for adduction–abduction and internal–external rotation, with no significant between-group difference in flexion–extension angular speed.

Finally, the Hotelling *T*^2^ test for knee joint moment indicated a significant difference between ACLR and control group at 57.52% to 61.28% (*t*\* = 17.52, *t*_*max*_ = 20.87, *p* = 0.025) and 74.41% to 77.75% (*t*\* = 17.58, *t*_*max*_ = 20.85, *p* = 0.028) of the stride cycle. The Bonferroni adjusted post hoc test indicated that the ACLR group had a smaller knee flexion moment than the control group at 57.43% to 62.18% (*t*\* = −3.61, *t*_*max*_ = −4.14, *p* = 0.005, *d*_*range*_ = 0.83 − 0.89) of the stride cycle (fig. 6).

**Figure 6:**
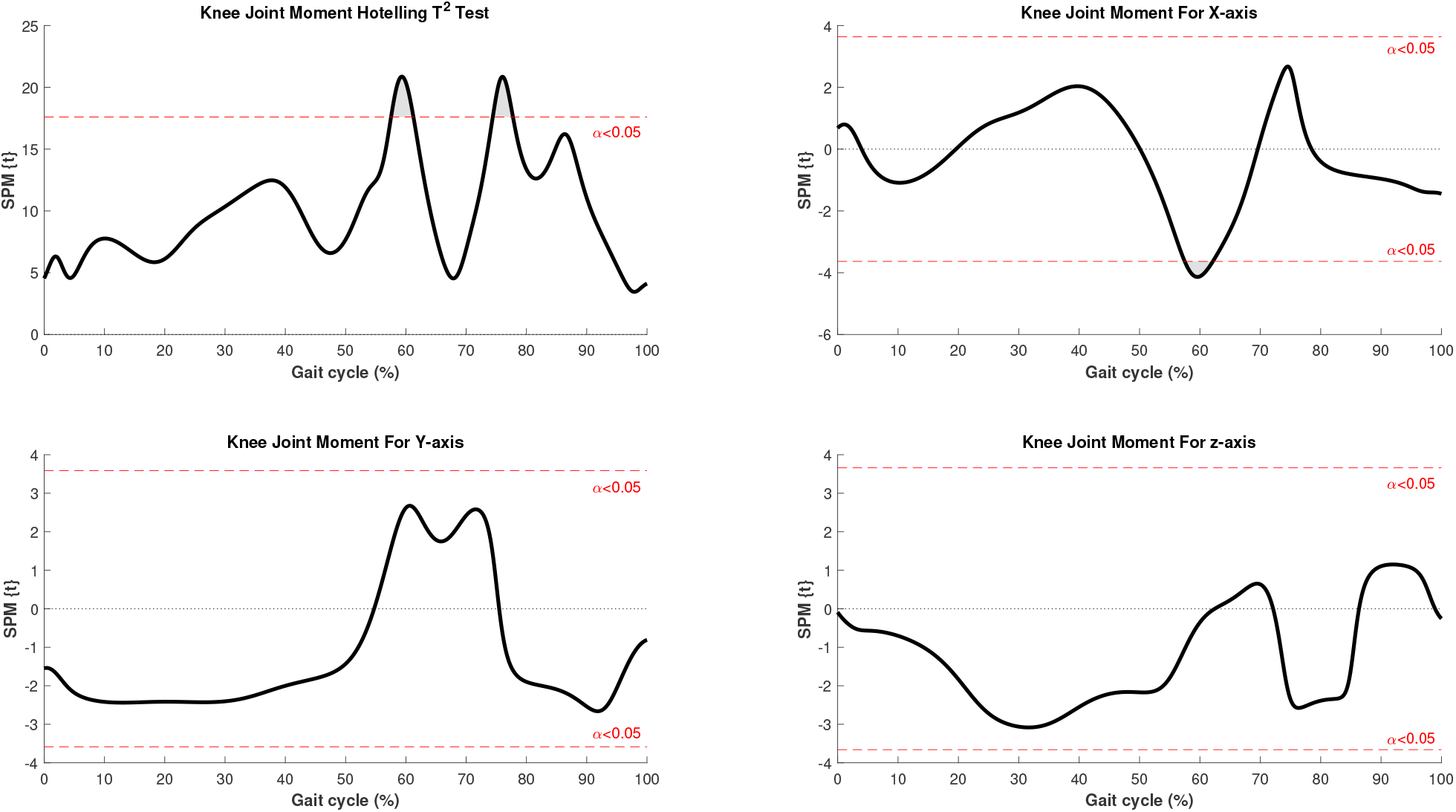
Reduced Knee Flexion-Extension Joint Moment in the ACLR Limb During a Specific Phase of the Gait Cycle. Statistical parametric mapping (SPM) analysis of knee joint moment. The upper-left panel presents the SPM{*T*^2^} trajectory from a Hotelling’s *T*^2^ test evaluating the three-dimensional knee joint moment simultaneously. The remaining panels present post hoc SPM{t} analyses of the sagittal (flexion–extension, X), frontal (adduction–abduction, Y), and transverse (internal–external rotation, Z) components. All analyses were performed using a two-tailed configuration. For visualization, the plots were scaled to highlight regions in which statistically significant between-group differences occurred. Internal knee joint moments were derived using inverse dynamics and normalized to body weight and height. The ACLR limb exhibited lower flexion–extension knee joint moments, with no significant between-group differences in adduction–abduction or internal–external rotation moments.

No statistically significant difference was found between the ACLR and control groups for knee adduction–abduction and internal–external rotation moments.

## 4. Discussion

The increase in muscle coactivation about the knee is a known neuromuscular adaptation following ACLR and has been implicated in altered joint-loading mechanisms that precede osteoarthritis. ^8,15^ However, the existing literature has studied this phenomenon as a discrete, phase-averaged quantity most often derived from the quadriceps and hamstrings, obscuring both the precise timing of within-stride differences and the contributions of other muscles that cross the knee. ^13,30^ As a result, it is unclear when during the gait cycle coactivation diverges between reconstructed and healthy joints, and how that divergence relates to concurrent joint mechanics. The present study aimed to address this by applying a continuous, time-varying, multi-muscle coactivation function (TMCf) across the entire gait cycle in ACLR individuals and matched healthy controls, and to concurrently evaluate this alongside knee joint angle, angular instantaneous speed, and moment to temporally characterize coactivation and joint loading.

The central finding of this study is that differences in coactivation between reconstructed and healthy limbs were concentrated within specific phases of the gait cycle. The absence of significant difference in CoM displacement and instantaneous speed suggest that the observed differences in knee coactivation and knee joint dynamics were not attributable to differences in walking speed or stride mechanics. Rather, these findings may reflect neuromuscular alterations associated with the reconstructed limb following ACLR (Figure 1). Significantly greater coactivation in the ACLR limb was found during most of the first 8% of the cycle, briefly around 29%, around 35–41%, and intermittently throughout the last 30%. These percentages respectively correspond approximately to the loading response, the late mid-stance phase, the terminal stance and early push-off phases, and intermittently throughout swing. ^38^ Contextualizing this with the functions and demands of each phase, this pattern is mechanistically coherent. The loading response is when the limb absorbs impact and the knee normally flexes to attenuate the ground reaction force. Heightened coactivation here may restrict knee flexion excursion, limiting the capacity of the joint to attenuate load through muscular work and redistributing mechanical stress from the reconstructed ligament onto passive structures such as the cartilage and meniscus. ^8,15^ During terminal stance and early push-off, elevated coactivation coincided with a difference in knee joint angle in knee internal–external rotation angle, which was present across this portion of the stride. These observations in conjunction may increase the stress borne by the cartilage and meniscus, as has been reported previously. ^15^ Approaching the transition into early initial swing, elevated coactivation coincided with a reduced knee flexion–extension moment. Increased coactivation raises active stiffness about the knee, and this in conjunction with the reduced joint moment may reflect a strategy that limits loading of the reconstructed ligament during propulsion. Together, these observations support the interpretation that coactivation after ACLR reflects a strategy of joint protection achieved through restricted knee motion. ^15,26^

These findings are consistent with prior work reporting greater quadriceps–hamstrings coactivation in the ACLR limb during the preparatory and heel strike phases. ^8^ However, because that study averaged coactivation within predefined phases, it could not identify where within those phases the difference occurred. The continuous analysis used here localizes the difference to the first 8% of the gait cycle. Additionally, our results agree with reports of elevated hamstrings coactivation during isometric tasks, suggesting that the increase observed in this study while walking is not attributable to the task alone but is a general feature of the reconstructed knee. ^9,10^

With regards to the joint kinematics and kinetics, the reconstructed knee exhibited a smaller knee flexion–extension moment than controls during early initial swing (57.43 – 62.18%). ^38^ Additionally, the reconstructed knee showed an altered knee internal–external rotation angle across nearly the entire stride and greater knee angular speed over most of the cycle, rather than the narrow late-stance window in which the moment differences were observed. This difference in distribution suggests that the reconstructed knee moves differently throughout gait while the net knee moment differs from controls only during propulsion. The joint-moment differences also occupied a different portion of the cycle than the coactivation differences, which were concentrated earlier, during loading response and terminal stance, and later, during swing. Elevated coactivation and reduced knee flexion–extension moment therefore emerged in distinct phases of gait. A reduced knee moment during the propulsive phase is a documented pattern after ACLR and is consistent with offloading or underutilization of the reconstructed knee, which can affect long-term joint health. ^39^ This is similarly supported by cohort evidence associating decreased knee joint loading with early osteoarthritis after ACLR. ^34^ Whether the earlier increased coactivation contributes to the later alteration in joint moment would require further continuous association analysis with sufficient statistical power.

The SPM analysis of a continuous, multi-muscle coactivation helps to bypass the discrete-time-point limitation of assessing coactivation. ^13,37^ Time localization of the increased coactivation to weight acceptance and push-off identifies specific windows of the gait cycle that could be a target for phase-specific rehabilitation rather than by generalized strengthening alone, which may improve the efficiency of rehabilitation regimen. Since the reducing hamstrings coactivation have been shown to lower tibiofemoral compressive force and joint stiffness in the reconstructed knee, ^10,27,28^ investigating more localized rehabilitation regimen may efficiently reduce the risk of knee osteoarthritis which require further investigation.

### 4.1 Limitations

Although current study used a novel statistical approach to characterize time-varying multi-muscle function and assess coactivation about the knee joint, several limitations should be considered when interpretating these findings. First, the sample included ten participants per group. Although the relatively large effect sizes observed for the coactivation differences suggest that the magnitude of the between-group differences was substantial, the modest sample size may limit the generalizability of these findings and increases the potential for sampling variability. Replication in larger cohorts is therefore warranted. Second, coactivation does not fully represent the mechanical consequences of joint co-contraction, which depend on the magnitude of muscle forces and their respective moment arms about the joint. Nevertheless, coactivation provides a useful measure of simultaneous muscle activation and has been used as an indicator of the neuromuscular contribution to active joint stiffness. Third, coactivation estimates are sensitive to electrode placement. Electrodes were therefore positioned according to SENIAM recommendations to minimize variability associated with electrode placement and improve measurement consistency. Fourth, several significant swing-phase windows encompassed less than 1% of the gait cycle. Although these brief windows indicate statistically significant differences in coactivation, their short duration may limit their contribution to overall joint mechanics relative to longer-duration phases. Finally, the cross-sectional design precludes conclusions regarding causality or temporal relationships among coactivation, knee joint mechanics, and ACLR status. Future longitudinal studies are needed to determine whether the observed alterations in coactivation are consequences of ACLR, adaptations associated with rehabilitation and recovery, or persistent neuromuscular characteristics that contribute to altered knee mechanics.

## 5. Conclusion

In this study, muscle coactivation about the knee was elevated in the ACLR limb during specific phases of the gait cycle. The differences in coactivation were most prominent during weight acceptance and the transition from terminal stance into early initial swing. A reduction in the knee flexion–extension moment was identified in a distinct phase of the cycle, while knee internal–external rotation angle differed from healthy controls across much of the stride.

Together, these findings indicate that the neuromuscular and mechanical differences between the ACR and healthy limbs are localized to identifiable portions of the gait cycle rather than distributed uniformly across the stride. When considered alongside prior evidence that sagittal-plane loading deficits are among the most persistent and clinically relevant gait abnormalities after ACLR, ^39^ as well as the absence of significant between-group differences in walking speed and stride distance, the present findings support multi-muscle coactivation as a time-resolved neuromuscular feature of altered knee mechanics. These findings highlight the potential utility of phase-specific assessment of coactivation during gait and may inform rehabilitation strategies targeting intervals in which altered neuromuscular coordination is most pronounced. Future longitudinal studies should confirm these patterns in larger samples and determine whether phase-specific coactivation is associated with altered knee joint loading and whether these alterations change with rehabilitation and recovery.

## Data Availability

All data produced in the present study are available upon reasonable request to the authors

## 6. Acknowledgement

All authors declare no conflict of interest. The results of the study are presented clearly, honestly, and without fabrication, falsification, or inappropriate data manipulation.

